# Summaries, Analysis and Simulations of COVID-19 Epidemics in Shanghai

**DOI:** 10.1101/2022.01.11.22269050

**Authors:** Lequan Min

## Abstract

Shanghai is the best city to prevent the spread of COVID-19 infection in China. Since February 20, 2020, Shanghai has experienced five waves of COVID-19. Out of a total of 388 patients with COVID-19 symptoms, 381 were cured and seven died. Medical staff achieved zero infection. This paper summarizes, analyzes and simulates COVID-19 epidemic in Shanghai. The simulation results show that for five waves of epidemics, after reaching the infection turning point, the blocking rate of symptomatic infection is over 99%. The administration needs to maintain the prevention and control implemented seven days after reaching the infection turning point until the new infection goes away.

## 1 Introduction

Over the past two years, the 2019 coronavirus disease (COVID-19) has placed tremendous pressures onto the prevention, control, and healthcare systems worldwide. Many countries have experienced multiple outbreaks of the COVID-19 epidemic due to incomplete preventive measures and virus variants. As of January 8, 2022, there are more than 298.9 million confirmed cases of COVID-19 with just under 5.5 million deaths globally (https://www.who.int/emergencies/diseases/novel-coronavirus-2019).

Shanghai is the best city to prevent the spread of COVID-19 in China. this paper summarizes, analyzes the five waves of the COVID-19 epidemic in Shanghai, estimates infection transmission rates, infection blocking rates, and preventive measures through modeling and numerical simulations.

## 2 Summary of Five Wave COVID-19 Epidemics in Shanghai

The dataset of the Shanghai COVID-19 epidemic was collected and edited from the official website [1]. Since 2020, there have been five waves of epidemics in Shanghai (more than three infected people). The Shanghai epidemics are summarized as follows:

1. *The first wave epidemic*. From January 20 to May 2, 2020, there were 339 infected people, seven of whom died. There were no new infections after 63 days. The highest number of people hospitalized with symptoms was 255 (days 23-24). The largest daily increase in infections was 27 on day 10. The first person recovered on day 4. Up to 34 people recovered on day 26. All hospitalized patients recovered after day 103.
2. During the period from May 3 to July 19, 2020, on May 17, May 20 and June 29, three imported symptomatic cases from other provinces were reported, and they were recovered on May 27, May 31, and July 19, respectively. No new infections reported from July 19 to November 8, 2020.
3. *The second wave epidemic*. From November 9, 2020 to January 2, 2021, cumulative seven people were infected and no deaths were reported. There were no new infections after the 14th day. The highest number of people hospitalized with symptoms was 7 (on days 14-19). The largest daily increase in infections was on days 11 and 13, with two infections. The first patient recovered on day 20. Up to 2 individuals recovered on day 26. All hospitalized patients cleared after day 56.
4. No infections were reported between January 3 and January 19, 2021.
5. *The third wave epidemic*. From January 20 to March 17, 2021, cumulative 22 people were infected and no deaths were reported. There were no new infections after the 16th day. The highest number of people hospitalized with symptoms was 22 (on days 14-16). The largest daily increase in infections was on the 1st day, with 4 infections. The first two patients recovered on day 18. Up to 3 individuals recovered on day 35. All hospitalized patients cleared after day 55.
6. No new infections were reported between March 18 and August 1, 2021.
7. *The fourth wave epidemic*. From August 2, 2021 to September 28, 2021, cumulative 10 people were infected and no deaths were reported. There were no new infections after the 24th day. The highest number of people hospitalized with symptoms was 9 (on days 24-34). The largest daily increase in infections was on day 19, with 3 infections. The first patient recovered on day 18. Up to 2 individuals recovered on days 44 and 48. All hospitalized patients cleared after day 57.
8. No infections were reported between September 29 and November 24, 2021.
9. *The fifth wave epidemic*. From November 25, 2021 to January 4, 2022, cumulative 7 people were infected and no deaths were reported. There were no new infections after the 22th day. The highest number of people hospitalized with symptoms was 7 (on days 22-26). The largest daily increase in infections was on the 1st day, with 3 infections. The first patient recovered on day 27. There was one recovered individual on days 27, 31, 32, 34, 36, 37 and 40 respectively. All hospitalized patients cleared after day 40. There are still 8 asymptomatic individuals under investigations.

In summary, during January 20, 2020 to January 4, 2022, Shanghai reported 388 COVID-19 infected individuals, 381 individuals recovered and seven died. There are 8 asymptomatic individuals to be investigated.

## 3 Analysis and Simulations of COVID-19 Epidemics in Shanghai

The first Shanghai epidemic infected 399 individuals and lasted 103 days. However the following four epidemics infected 7, 22, 10, and 7 individuals, and lasted 56, 55, 57, and 40 days, respectively. The time of clearing new infections of the first Shanghai epidemic was 64 day. However the times of clearing up new infections of the four epidemics were 14 day, 15 day, 24 day, and 22 day, respectively. The reported data suggest that after the first epidemic, the Shanghai authority has implemented very effective prevention measures such that decreased largely infected individuals, healthcare expenditures and workloads.

Figures 1-3 show the outcomes of the current symptomatic individuals the cumulative recovered symptomatic individuals and the current covered individuals of the five wave epidemics in Shanghai. The three figures display visually the evolutions of the five epidemics in Shanghai.

**Figure 1.**
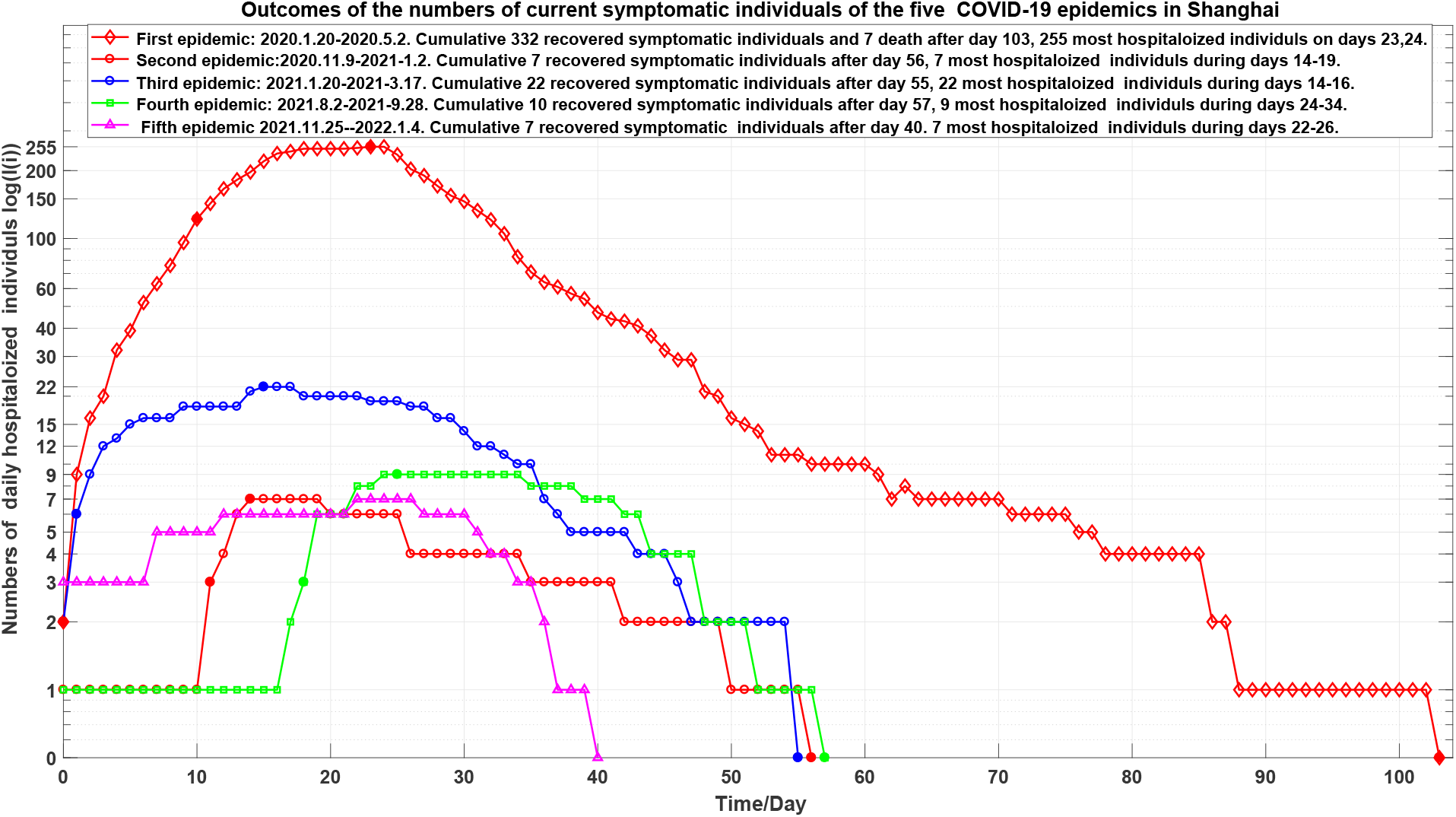
Outcomes of the current symptomatic individuals of the five wave epidemics in Shanghai. The first epidemic marked by 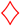. The second epidemic marked by 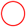. The third epidemic marked by 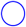. The fourth epidemic marked by 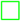. The fifth epidemic marked by 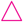.

**Figure 2.**
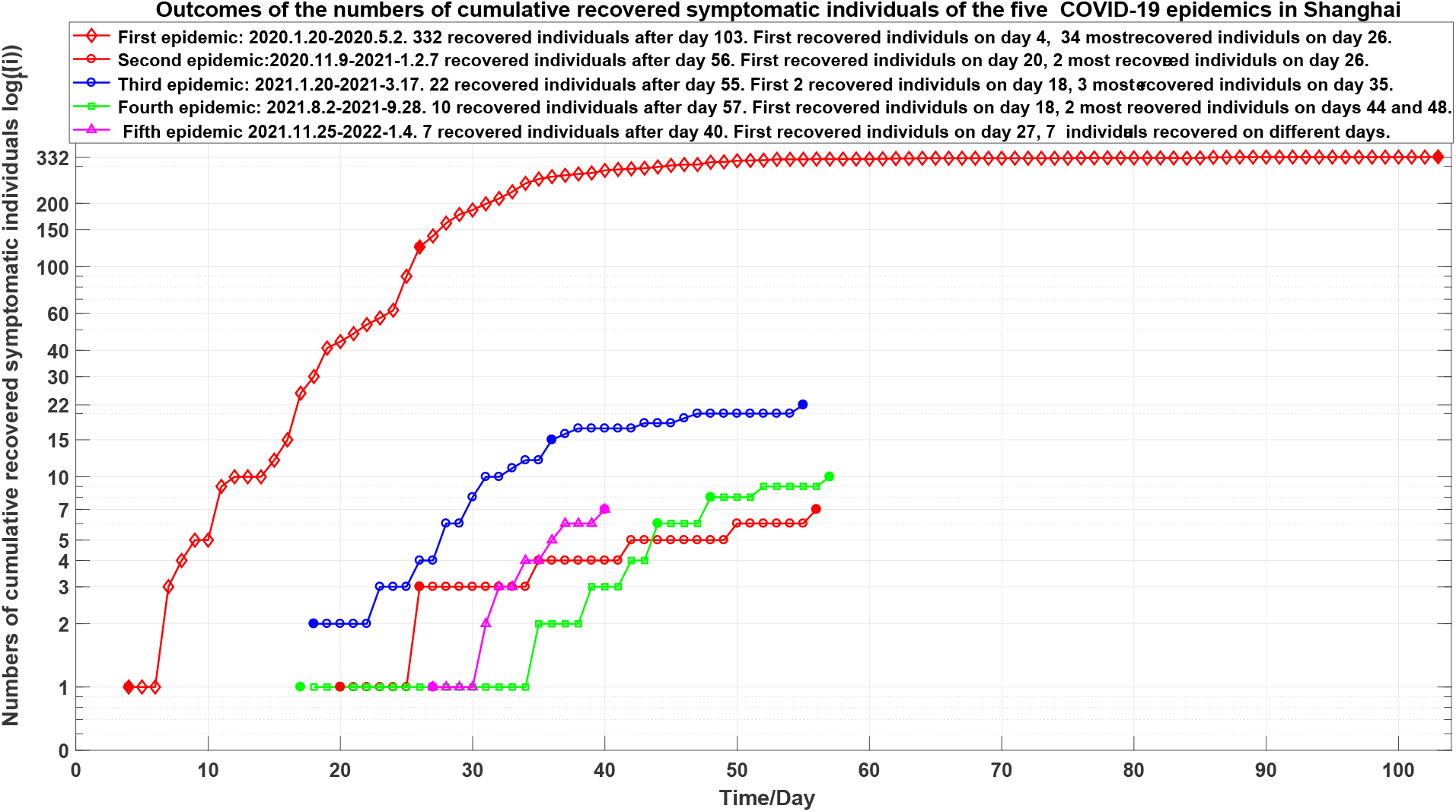
Outcomes of the cumulative recovered symptomatic individuals of the five wave epidemics in Shanghai. The first epidemic marked by 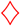. The second epidemic marked by 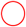. The third epidemic marked by 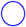. The fourth epidemic marked by 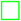. The fifth epidemic marked by 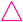.

**Figure 3.**
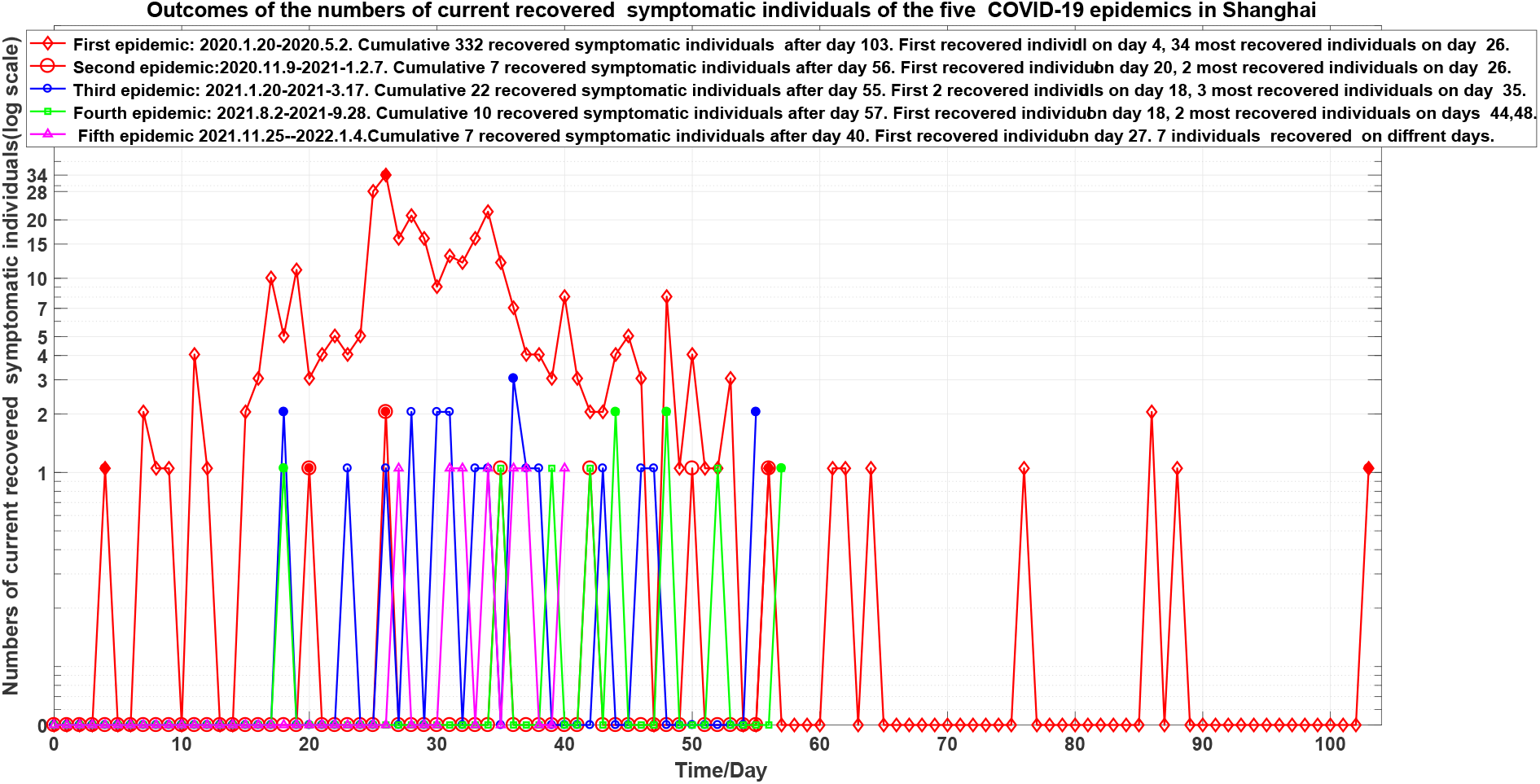
Outcomes of the current covered individuals of the five wave epidemics in Shanghai. The first epidemic marked by 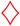. The second epidemic marked by 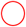. The third epidemic marked by 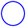. The fourth epidemic marked by 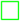. The fifth epidemic marked by 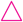.

In order to estimate numerically the transmission rates and blocking rates to symptomatic infections, we need to set up mathematic models to simulate the dynamics of spread of infection disease.

For the first epidemic, the symptomatic infected individuals (*I*) and the asymptomatic infected individuals (*I*_a_) infect the susceptible population (*S*) with the transmission rates of *β*_11_ and *β*_21_, respectively, making *S* become symptomatic infected individuals, and with the transmission rates of *β*_12_ and *β*_22_, respectively, making *S* become asymptomatic individuals. Then, a symptomatic individual is cured at a rate *κ*, an asymptomatic individual returns to normal at a rate *κ*_a_. An infected individual dies at a rate *α*. Here all parameters are positive numbers. Assume that the dynamics of an epidemic can be described by *m*-time intervals, which correspond different prevention and control measures, and medical effects. At *i*th time interval, the model has the form [2, 3]:

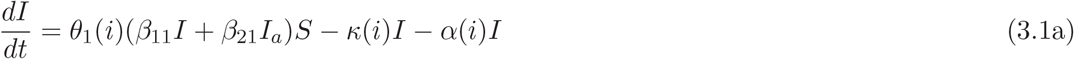

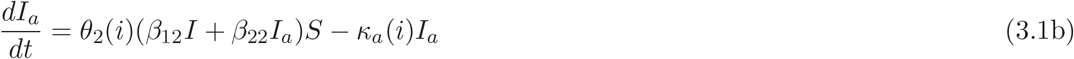

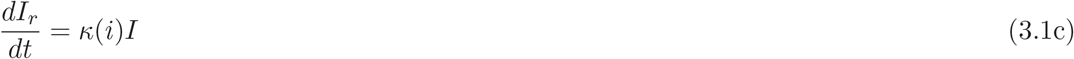

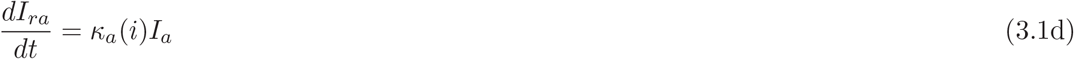

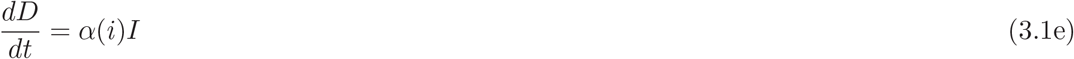

where Θ_1_(*i*) = (1 − *θ*_1_(*i*)) and Θ_2_(*i*) = (1 − *θ*_2_(*i*)) (*i* = 1, …, *m*) represent the blocking rates to symptomatic and asymptomatic infections, respectively.

For other epidemics, the model has simply the form

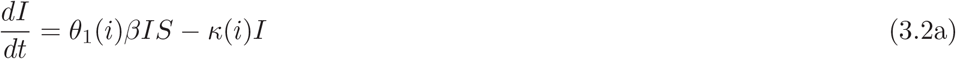

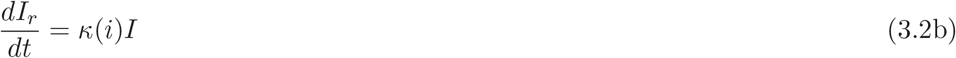

where Θ_1_(*i*) = 1 − *θ*_1_(*i*) represents the blocking rate to symptomatic infections, and *β* represents the transmission rate of the symptomatic individuals to make *S* become symptomatic individuals.

The parameters of equations (3.1) and (3.2) of the five wave epidemics of Shanghai are given in Table 1. The calculated transmission rates *β*, blocking rate 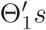 to symptomatic infections are listed in Table 2. It follows that:

**Table 1.**
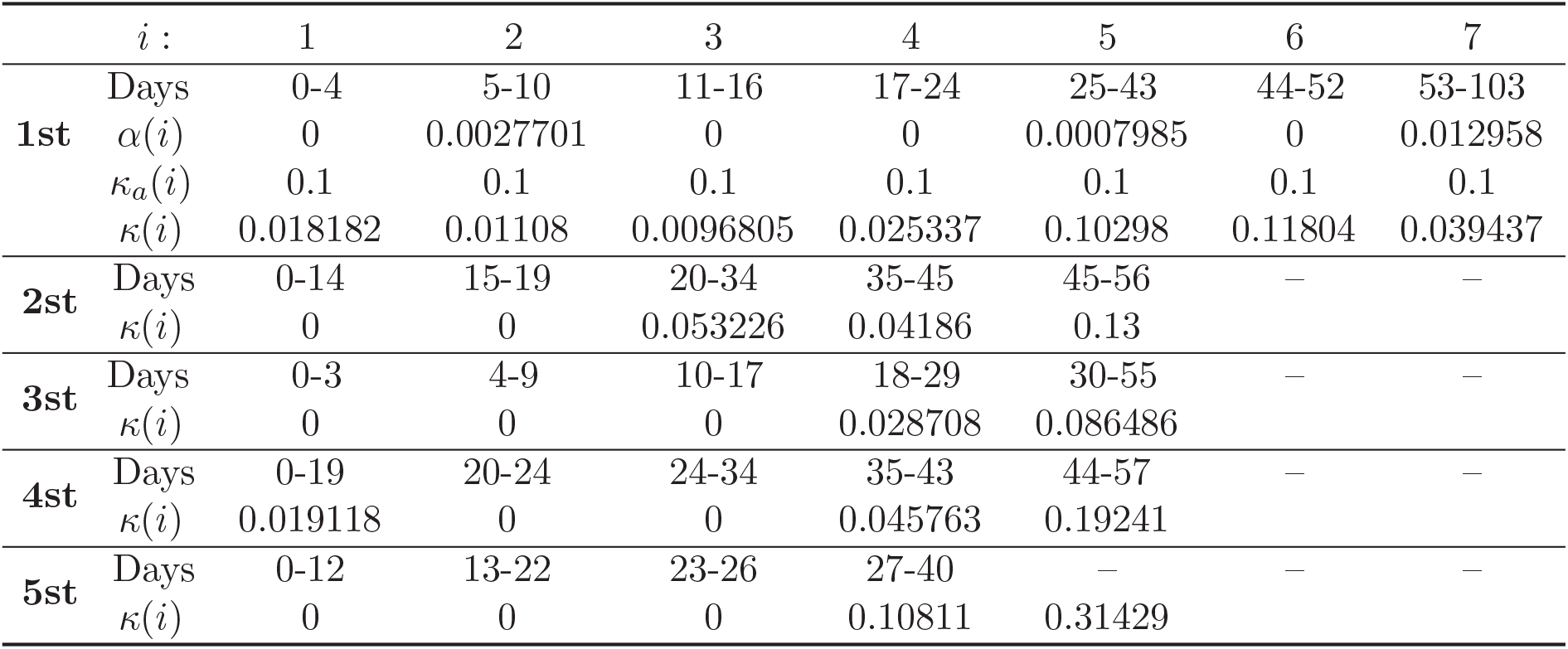
Equation parameters of the five wave epidemics of Shanghai during 2020-2021.

**Table 2.** Equation parameters of the five wave epidemics of Shanghai. For the first epidemic, *β*_21_ = 0.23, *β*_12_ = 0.39756, *β*_22_ = 0.014199.

| | $i :$ | 1 | 2 | 3 | 4 | 5 | 6 |
| --- | --- | --- | --- | --- | --- | --- | --- |
| **1st** $\beta_{11}$ | Days | 0-4 | 5-10 | 11-16 | 17-24 | 25-43 | 43-103 |
| 0.59643 | $\Theta_1(i)$ | 0 | 66.6% | 82.7% | 95.5% | 99.1 | 100% |
| | $\Theta_2(i)$ | 0 | 80% | 82.7% | 81.1% | 83.1 | 100% |
| **2st** $\beta$ | Days | 0-14 | 15-19 | 20-56 | – | – | – |
| 0.13881 | $\Theta_1(i)$ | 0 | 99.6% | 100% | – | – | – |
| **3st** $\beta$ | Days | 0-3 | 4-9 | 10-17 | 18-29 | 30-55 | – |
| 0.59546 | $\Theta_1(i)$ | 0 | 88.5% | 95.5% | 99.5% | 100% | – |
| **4st** $\beta$ | Days | 0-19 | 20-24 | 24-34 | 35-57 | – | – |
| 0.11352 | $\Theta_1(i)$ | 0 | 28.9% | 99.9% | 100% | – | – |
| **5st** $\beta$ | Days | 0-12 | 13-22 | 23-26 | 27-40 | – | – |
| 0.07934 | $\Theta_1(i)$ | 0 | 73% | 99% | 100% | – | – |

1. Transmission (Baseline) Rates for first and third outbreaks Epidemics are almost the same. However, higher blocking rates end the second outbreak 48 days earlier than the first.
2. The third, fourth, fourth and fifth epidemic (baseline) transmission rates were very low. This explains more about effective infection prevention (Example: more people carry masks in public places, less public places gatherings). The blockage rate 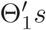 reached more than 99% after reaching the inflection returning point for the five epidemics (See Figure 1 and Table 2).
3. The blocking rate 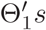 reached more than 99% after reaching the inflection turning point for the five epidemics (See Figure 1 and Table 2).
4. For the five epidemics, the blocking rates 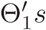 reached over 99% after the infection turning point days (see Figure 1 and Table 2).

Figures 1-3 show the outcomes of the current symptomatic individuals the cumulative recovered symptomatic individuals and the current covered individuals of the five wave epidemics in Shanghai. The three figures display visually the evolutions of the five wave epidemics in Shanghai.

Figures 4 and 5 show the simulation results of equations (3.1) and (3.2 for the outcomes of the current symptomatic individuals and the cumulative recovered symptomatic individuals. Observe that simply models can describe approximately the complex evolutions of the five wave epidemics in Shanghai. In particularly, we can give the estimations to the infection blocking rates with the help of the simulations.

**Figure 4.**
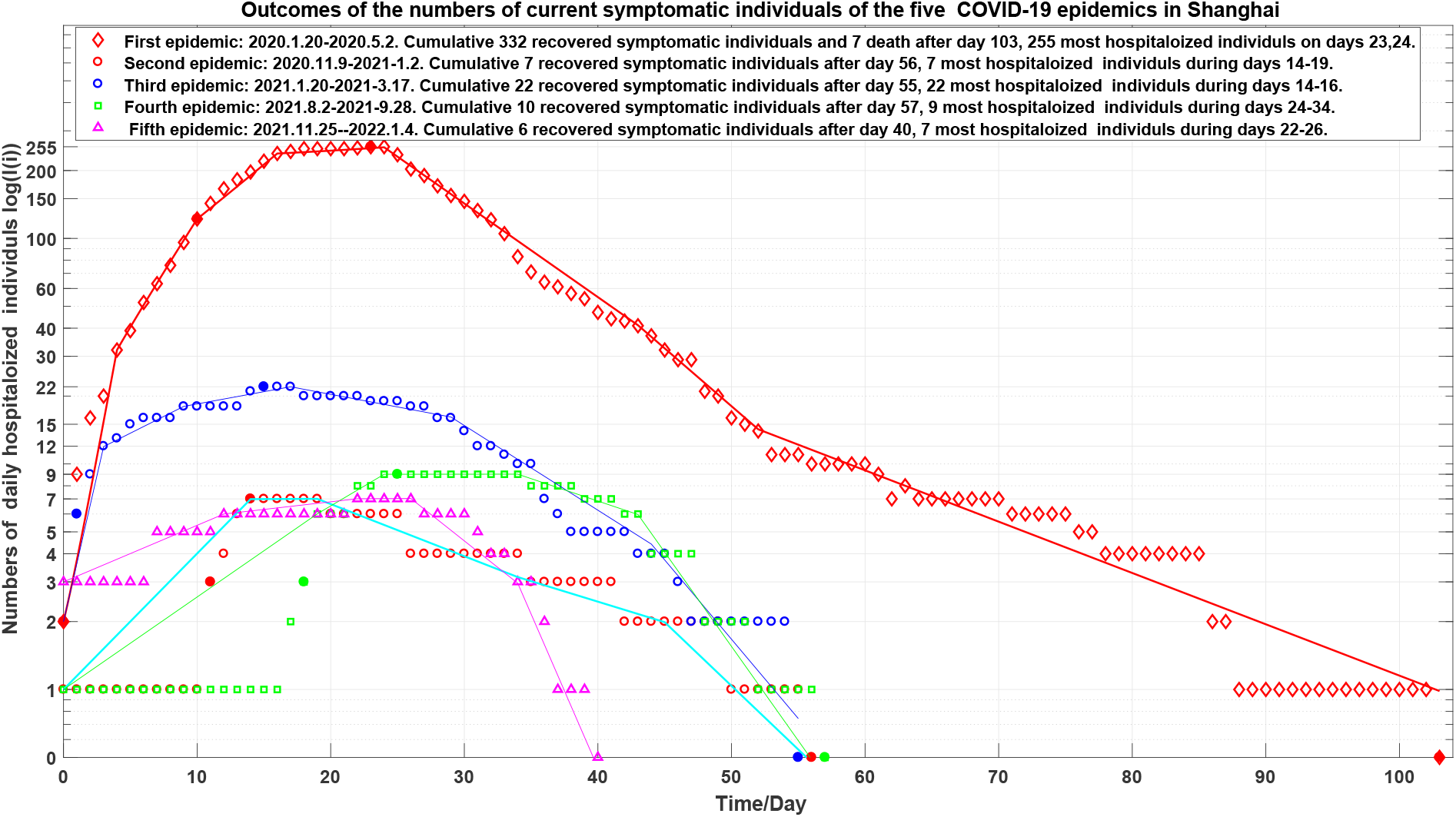
Outcomes of the current symptomatic individuals of the five wave epidemics in Shanghai marked by 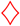, 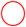, 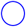, 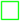 and 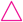, respectively. The corresponding simulations are displayed via solid lines colored by red, cyan, blue, green and magenta, respectively.

**Figure 5.**
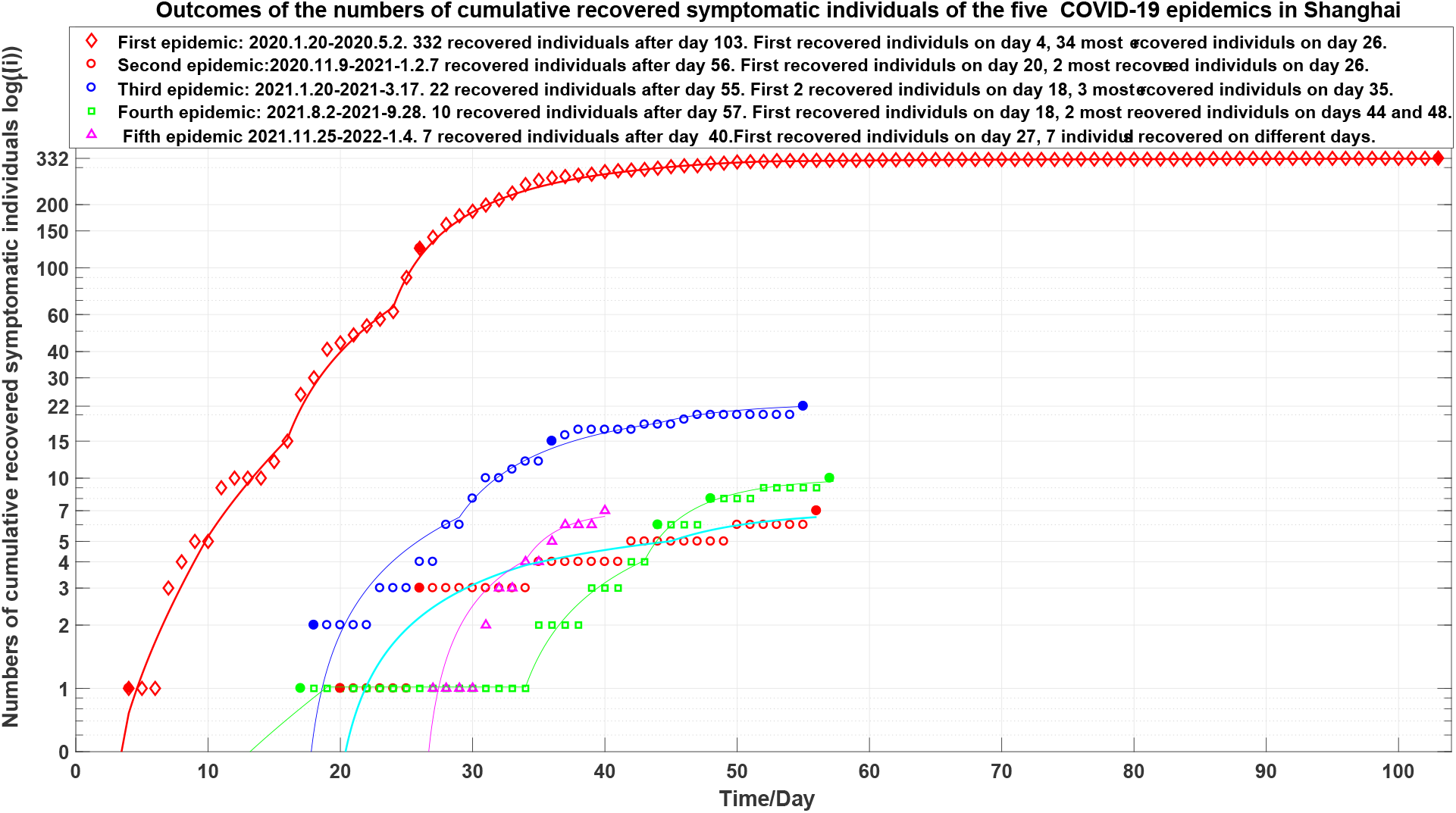
Outcomes of the cumulative recovered symptomatic individuals of the five wave epidemics in Shanghai marked by 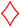, 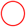, 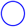, 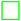 and 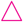, respectively. The corresponding simulations are displayed via solid lines colored by red, cyan, blue, green and magenta, respectively.

## 4 Conclusions

The main contributions of this paper are summarized as follows:

1. It is the first time to summary the five wave Shanghai epidemics. It shows a clear picture to prevent the spread of the Shanghai epidemics.
2. It introduces two models to simulate the Shanghai epidemics. The simulation results can provide reasonable interpretation and estimation of prevention and control measures and effectiveness of treatments.
3. It suggests that it needs a blocking rate of more than 95% to prevent the spreads of the COVID-19 epidemics.
4. It suggests that good prevention and control measures and treatments may end new COVID-19 infections in 40 days.

A recommendation to avoid multiple outbreaks of an epidemic is competent authorities should at least maintain preventive and control measures implemented 7 days after inflection turning point until all new infections have been cleared [3].

It is not wise strategy to withdraw all prevention and control measures before the number of the all infected people have been cleared. 100% blocking the speed at which COVID-19 infection spreads is key Strategies for early clearance or reduction of epidemic spread possible.

Strict prevention and control strategies implemented in Shanghai authority are not only effective but also necessary. It is expected that this research can provide better understanding, interpretation and leading the spread and control measures of epidemics.

## Data Availability

All data produced are available (need to edit) online at
http://wsjkw.sh.gov.cn/.

http://wsjkw.sh.gov.cn/.

## Data availability statement

Data are available on reasonable request. Please email the author.

## Ethical Statement

Not applicable/No human participants included.

